# Can identifying recent *Mycobacterium tuberculosis* infection support faster tuberculosis vaccine trials?

**DOI:** 10.64898/2026.10.19.26365122

**Authors:** Liza Hadley, Tyler S Brown, Kristin Nelson, Stephen M Kissler

**Affiliations:** University of Colorado Boulder, Boulder, CO, USA; University of Cambridge, Cambridge, UK; University of Colorado Anschutz School of Medicine, Aurora, CO, USA; Emory University, Atlanta, GA, USA

**Keywords:** Tuberculosis, vaccine, clinical trial, modelling, sample size, TASA, IGRA

## Abstract

**Background:** TB vaccine development is hampered by the natural history of M. tuberculosis: most infected people rarely progress to disease, so trials must be very large and long to accrue sufficient disease endpoints. Current trials enrol IGRA-positive participants (IGRA: Interferon-Gamma Release Assays), but IGRAs remain positive for many years and cannot distinguish recent from remote infection. Tests for recent infection are now in development, and we argue they could make trials substantially more efficient. Here, we investigate how screening participants prior to enrolment with a test for recent *M.tb* infection could impact the total number of enrollees required for an adequately-powered TB vaccine trial.

**Methods:** Using a statistical model of vaccine trial recruitment, we estimated the number needed to contact, test, and enrol to achieve an ‘effective’ trial (defined as 50 disease endpoints), under IGRA-gated enrolment (life-long duration of positivity) vs. enrolment with a test for recent infection. We considered five recruitment strategies: non-test-based enrolment for ages 15-49, ages 12-34, and household contacts of recent cases; and test-based enrolment using IGRA, and a test for recent infection with two-year duration of positivity. The model explored trial recruitment in South Africa, Kenya, and India.

**Results:** Compared with an extended-positivity test, a test for recent infection reduced the median number needed to enrol by 4.2- to 6-fold under favourable conditions. It also outperformed non-test-based strategies, including enrolling household contacts. This comes at the cost of substantially more testing, but crude estimates suggest recency-based enrolment would still be cheaper overall provided the new test does not cost more than roughly 5-10 times an IGRA.

**Conclusions:** Diagnostics for recent *M.tb* infection have the potential to improve vaccine trial efficiency, but more research is now needed to understand test characteristics and to evaluate their use in this setting. Our Viewpoint sets out the test attributes that would need to be established, such as time to positivity, duration of positivity, sensitivity, and specificity, before such tests could be used in vaccine trials.

## Main text

The natural history of tuberculosis (TB) has made it challenging to develop new, effective TB vaccines [1]. Following *Mycobacterium tuberculosis* (*M.tb*) infection, only 5-10% of individuals will develop TB in their lifetimes, with some individuals undergoing rapid progression to disease within 1-2 years (‘fast progressors’) and the majority experiencing a low annual chance of progressing to disease [2]. Consequently, trials must be large and long to gather sufficient disease endpoints for evaluating a vaccine. To address this challenge, TB vaccine trials have used test-gated enrolment, where participation is limited to individuals with positive interferon gamma release assay (IGRA) results and no evidence of active disease. This strategy enriches trials with participants with prior *M.tb* infection and evaluates whether a vaccine prevents progression to TB [3]. However, because only a small percentage of IGRA-positive participants will develop TB over the duration of most vaccine efficacy studies, thousands of individuals must still be enrolled to achieve a sufficiently-powered trial (e.g. [4]).

Diagnostics that can identify *M.tb*-infected participants at risk for more imminent progression to disease could improve test-gated enrollment strategies, allowing a smaller number of participants to be followed for progression to active TB and leading to faster and more efficient trials. Candidate diagnostics for this purpose include tests that can distinguish recent versus distant (i.e. acquired longer ago) *M.tb* infection [5,6], given that risk of progression to TB is substantially higher among individuals in the first 1-2 years after infection. IGRAs have an extended, often life-long, duration of positivity (DOP) and thus cannot distinguish recent versus remote *M.tb* infection. Tests for recent infection have inherently shorter DOP, such that they revert to negative after some shorter interval after infection. Here, we investigate how screening candidate participants prior to enrolment with a test for recent *M.tb* infection could impact the total number of enrollees required for an adequately powered TB vaccine trial.

Using a statistical model of vaccine trial recruitment, we estimated the number needed to contact, test, and enrol to achieve an ‘effective’ trial, under IGRA-gated enrolment (life-long DOP) versus enrolment based on a test for recent infection, with a short DOP. An ‘effective’ trial is defined here as continually recruiting until at least 50 disease endpoints occur - similar to endpoint accrual for past Phase 2b trials [4]. Individuals in our model were randomly sampled from synthetic populations with age distributions matched to South Africa, Kenya, and India [4,7]. Individuals were enrolled if they met three requirements: aged between 15–49 years; disease-free; and (for test-based strategies) tested positive for *M.tb* infection in the past σ years. While current trials include individuals 15 and older, recruiting younger ages is desirable because it targets individuals as they enter a period of high infection and TB risk, so we also consider a trial which recruits children as young as 12 years of age [8]. For simplicity, we assumed perfect test specificity and sensitivity and immediate positivity upon infection. We also considered another method of enriching trials for recently infected individuals: recruiting household contacts of TB cases [9].

We considered five recruitment strategies: non-test-based enrolment for ages 15-49 years (*no test: standard*), ages 12-34 years (*no test: young*), and household contacts of recent cases (*no test: household*); and test-based enrolment using IGRA (*test: IGRA*), and a test for recent infection with a two-year DOP (*test: recent infection*) (Figure 1). From Figure 1, we see that trial recruitment using a test for recent infection yielded a 4.2-to 6-fold median reduction in the number needed to enrol across the three countries we considered, compared to IGRA-gated enrollment. Compared to non-test-based strategies, median reductions in number needed to enrol for the recent infection test-based strategy spanned from 8.5-to 16.3-fold for the “*no test: standard*” strategy, 8.9-to 19.4-fold for the “*no test: young*” strategy, and 2.3-to 4.1-fold for household-based enrollment.

**Figure 1:**
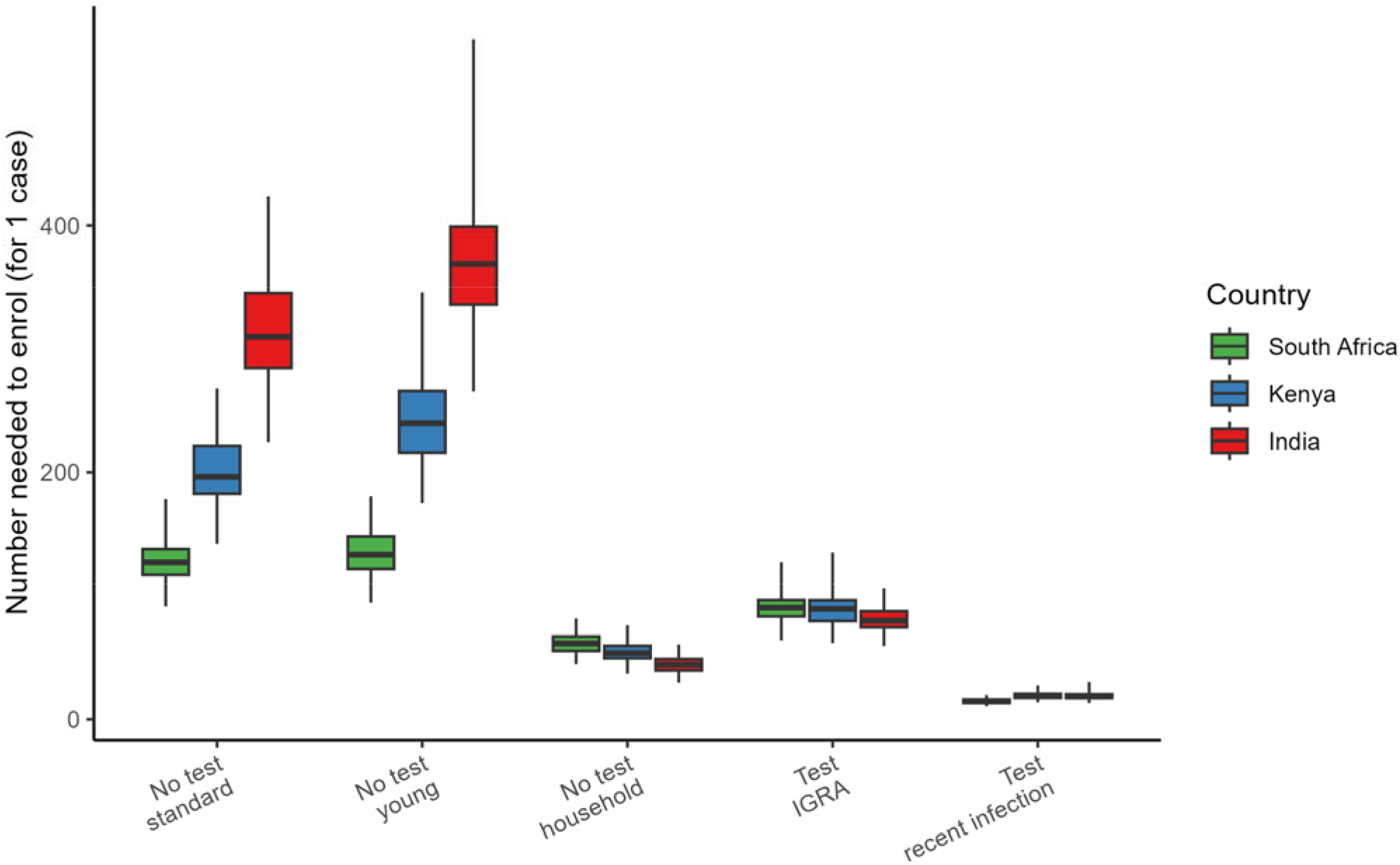
Estimated number needed to enroll to gather one trial endpoint, across countries and recruitment strategies. The five recruitment strategies shown in order are: non-test-based enrolment for standard ages 15-49y, younger ages 12-34y, and household contacts of ecent cases; test-based enrolment using an infection test with extended duration of positivity (IGRA), and two years duration of positivity (recent infection). Box plots depict the mean, interquartile range, and full range of number enrolled across 100 simulations, under baseline parameter values. The model here uses a 4-14% age-varying probability of being a ‘fast progressor’ for India and Kenya, and 4-19% for South Africa. See Supplementary Table S2 for a description of each scenario. Raw numbers corresponding to this plot are reported in Supplementary Table S4 and S5.

Figure 1 presents estimates of the number needed to enrol only. When using a test for recent infection over IGRA, the total number needed to test would increase substantially. Our estimates suggest that approximately ∼600 individuals would need to be tested per disease endpoint using a recency test with a 2-year duration of positivity, compared to only ∼130 for IGRA, in a high-burden South Africa-like setting (Supplementary Figure S4). Kenya and India estimates showed similar 3.4-to 3.7-fold increases in testing volume (Supplementary Figure S6). This added testing volume would appear to raise costs, but is counterbalanced by the corresponding 4.2 to 6-fold reduction in the number of individuals enrolled in the trial. Under the assumption that trial participation for a single enrolled individual would be at least 150 times more expensive than a single IGRA, our estimates suggest that recency test-based enrolment would be cheaper overall provided a recency test costs no more than 5-10 times an IGRA (Supplementary Table S4).

We build on the work of Cobelens et al. which focused on IGRA only [10]. Our results on required sample sizes are consistent with those of Cobelens et al., which estimated that at 2% annual risk of TB infection (ARTI) (e.g. Kenya), a no-test trial would need ∼124% more enrolled individuals than a trial using IGRA-based enrolment, while at 4-6% ARTI (e.g. South Africa), a no-test trial would need only be ∼8-36% larger [10].

Our analysis has limitations. First, we assume perfect test sensitivity and specificity, as test performance characteristics are ill-defined for both technologies: for IGRA, there is no agreed reference standard (comparator) against which to establish true sensitivity and specificity; and for recency tests, these are an emerging class for which validation is ongoing. Assuming perfect test performance in our model isolates the effect of duration of positivity, the parameter central to our comparison. Since our key outputs are relative comparisons between strategies, a degree of imperfect sensitivity/specificity would likely affect each strategy proportionately. Second, our model used a test for recent *M.tb* infection with a 2-year DOP, but this parameter is not yet well-described. Better quantifying DOP in candidate diagnostics in this class would help inform their potential use in TB vaccine trials. A third limitation is significant remaining uncertainty about how well TB vaccines might perform across pre-disease states. Vaccine trials gated using recency tests, undertaken with the goal of expediting vaccine evaluation, could yield negative or difficult to interpret results if test positivity correlates with more advanced, pre-disease states in which vaccination may be less effective. Further work should consider the trial stages at which these techniques may be appropriate to implement and further characterize the immunological profile of persons with recent *M.tb* infection based on these new assays. Optimizing trials for efficiency, while retaining the ability to clearly evaluate efficacy across at-risk populations, remains a central priority for the ongoing and future evaluation of candidate TB vaccines. Our results suggest that diagnostics for recent *M.tb* infection are one potential tool for improving the efficiency of these trials, but future work is needed to better evaluate their use and possible limitations in this context.

## Acknowledgements

We are grateful to Munyaradzi Musvosvi and Mark Hatherhill for their feedback on draft versions of this manuscript.

TSB and SMK are grateful for the joint CU Anschutz-CU Boulder AB-NEXUS grant that supported this work. SMK is also supported by a Mathematical Modeling of Policy Options for Evolving Public Health Challenges award from the U.S. National Science Foundation and Centers for Disease Control and Prevention (DMS-2436340), and TSB is supported by a K08 award from the National Institute of Allergy and Infectious Diseases (K08AI166125). KNN is supported by NIH/NIAID K01AI166093.

We acknowledge the use of AI (Claude Code) to support the development of model code and improve code efficiency, including generating a workflow for fine-detail model fitting of the force of infection.

## Author contributions

SMK, KN, and TSB posed the research question. All authors contributed to the design of this study. LH and SMK developed the model code and performed all analyses. LH drafted the first version of the manuscript. All authors provided critical feedback on analyses and of the manuscript, and approved the final version for publication.

## Data availability

All data and model code used in this article are available on GitHub at https://github.com/skissler/MINA_TB.

