## Supplementary Material for "Can identifying recent *Mycobacterium tuberculosis* infection support faster tuberculosis vaccine trials?"

#### Supplementary methods

##### Model overview

We developed a statistical model of vaccine trial *recruitment*, using both analytic and stochastic approaches to improve credibility. The main text shows results of the stochastic approach. In both cases, our objective function is to achieve an 'effective' trial, defined here as continually recruiting until at least 50 disease endpoints[1] have been enrolled. This value is motivated by the number of disease endpoints observed in the recent M72/AS01E phase IIb trial by Tait et al. [3]. Individuals are sampled from the population aged 15-49 years, with a constant South Africa-like population age distribution (UN WPP 2024 estimates) [3,4]. Requirements for enrolment in the model are as follows: Disease-free (as the vaccine is given prior to disease in current trials); AND Tests positive for *M.tb* infection (to increase the chance of progressing to disease during a three year trial). For simplicity, our model assumes perfect test specificity and sensitivity and immediate positivity upon infection. Note there is no vaccination in the model; we simulate vaccine trial *recruitment* only.

The analytic approach uses fundamental probability theory to estimate e.g. P(disease-free & positive & disease endpoint | age a), P(positive & disease endpoint | disease-free & age a), and P(disease endpoint | positive & disease-free & age a). From these, we can calculate the expected number needed to contact, test, and enroll for an effective trial (50 disease endpoints). See 'Detailed model equations' below for a full derivation. The stochastic approach on the other hand simulates random draws from probability distributions, following the below algorithm:

```
while (n_cases < 50) {
        Draw a person of random eligible age from the population.
        Draw their progression type (slow/ fast).
        Draw their time to infection tinf.
        Draw their primary reversion status (reverted/still positive).
        Draw their time to symptoms tsymp.
        If tsymp < age, the person is disease-free so we test them. Add 1 to n_tested.
        If tinf < age & still positive, the person is infected and has not reverted. They test positive so
        enrol them. Add 1 to n_enrolled.
        If fast, add 1 to n_fast. If slow, add 1 to n_slow.
        Update expected n_cases.   }
```

In both approaches, we additionally consider how long ago individuals were infected, to explore different *M.tb* infection tests and their duration of positivity. Duration of positivity in the model ranges from 6 months (i.e. only recently infected individuals would test positive) to essentially lifelong (ever-infected

---

[1] When infected with *M. tuberculosis*, a proportion of individuals appear to progress to disease within the first few years of being infected (here termed 'fast progressors') while others can take many years to progress or will never contract disease (here termed 'slow progressors') [1]. It is unclear what factors make an individual more or less likely to progress to disease at a given rate, but it is thought that the probability of being a 'fast progressor' increases with age of infection [2]. We consider a disease endpoint as an individual (fast or slow) who progressed to disease *during the trial*.

individuals would test positive). The extended duration of positivity represents IGRA-like infection tests, while the short duration of positivity aims to explore the potential of new 'recency' tests in vaccine trial design, such as the TB T-cell Antigen-Specific Activation assay [5–7]. Primary reversion (after first infection) is considered in the stochastic approach; the analytic approach assumes no reversion. The different infection windows and test and enrolment outcomes considered in our model are depicted in Supplementary Figure S1.

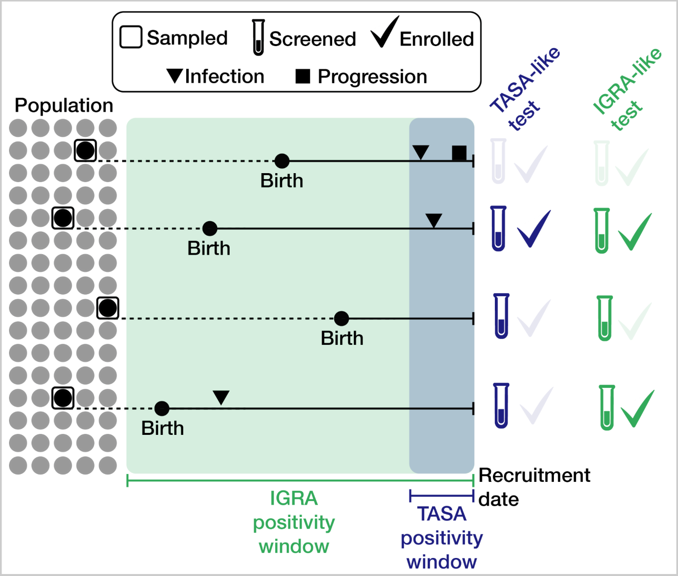


***Supplementary Figure S1:* A depiction of model enrolment.** *Individuals are drawn at random from the population of eligible ages, one at a time. Their time to infection, time to symptoms, and progression type (fast/slow) are drawn from probability distributions. If an individual is disease-free AND infected in the IGRA/TASA positivity window (controlled by parameter σ), we enrol them. We continue to contact, test, and enrol until a total of 50 disease endpoints have been enrolled.*

#### Detailed model equations

##### Analytic approach

The analytic approach estimates the number of people that need to be contacted, tested, and enrolled in order to enrol $\mathrm{n}^*$ disease endpoints in expectation in different settings. Here a disease endpoint is an individual who develops disease during the course of a 3-year trial. The analytic approach does not model reversion. For simplicity, let us count fast and slow progressors separately. For those individuals who are positive but disease-free on recruitment, we assume that almost all fast progressors would develop disease during the trial (calculated as $P(X \leq 3)$ for $X \sim Exp(1.5)$, i.e. $1 - e^{-1.5\times3} \approx 99\%$ ). Similarly, we assume a proportion $1 - e^{-0.003\times3} \approx 1\%$ of slow progressors would develop disease during the trial.

In the contact step, the aim is to identify individuals who are disease-free AND infected in past σ years ('positive') AND will become a disease endpoint. Consider fast progressors first. For this, we utilise the following probability expression conditioned on an individual of age a:

$$P(\text{disease-free \& positive \& fast} \mid \text{age a}) \;=\; p_f \int_{t=a-\sigma}^{a} \left(\rho e^{-\rho t} e^{-\mu_f(a-t)}\right) dt \qquad (1)$$

where $p_f$ is the baseline probability of being a fast progressor, $\rho e^{-\rho t}$ is the probability of getting infected at time t given constant force of infection ρ, and $e^{-\mu_f(a-t)}$ is the probability of not developing active TB disease by age a after getting infected at age t. Fast progressors progress to active TB disease at a rate $\mu_f$.

Integrating the equation and assuming our population has age distribution $f_a(a)$, we produce the following *unconditional* probability expression for the contacting step for fast progressors:

$$P_1 \;:=\; P(\text{disease-free \& positive \& fast}) \;=\; \int_a (p_f \rho \,\frac{e^{-\rho(a-\sigma)}\; e^{-\mu_f \sigma} \;-\; e^{-\rho a}}{\rho - \mu_f} f_a(a))\, da \quad (2)$$

Adding in the proportion of slow progressors who become disease endpoints, we have that for the contacting step the total probability of an individual meeting all criteria is:

$$P(\text{disease-free \& positive \& disease endpoint}) \;=\; (1 - e^{-3\mu_f})P_1 \;+\; (1 - e^{-3\mu_s})P_1' \quad (3)$$

where $P_i'$ is analogous to $P_i$ but with fast and slow-progressor parameters swapped (i.e. $p_f \rightarrow p_s$ and $\mu_f \rightarrow \mu_s$, and vice versa).

In the testing step, individuals need to be infected in past σ years ('positive') AND will become a disease endpoint GIVEN they are disease-free. Note that our model assumes tests have perfect specificity and sensitivity to detect infection, no reversion, and immediate positivity upon infection. Focusing on fast progressors and utilising the definition of conditional probability, we can say that:

$$P(\text{positive \& fast} \mid \text{disease-free \& age a}) \;=\; \frac{P(\text{disease-free \& positive \& fast} \mid \text{age a})}{P(\text{disease-free} \mid \text{age a})} \;. \quad (4)$$

The numerator has been calculated in equation (1). The denominator can be expressed using the law of total probability (with terms 'disease-free & ever infected & fast', 'disease-free & ever infected & slow', and 'never infected', given age a respectively). After algebra using similar ideas to equation (1) and integrating over age as before, we produce the following unconditional probability expression for the testing step for fast progressors:

$$P_2 \;:=\; P(\text{positive \& fast} \mid \text{disease-free}) \;=\; P_1 \;/\; \int_a (\sum_{i=f,s} [p_i \rho \frac{e^{-\mu_i a} \;-\; e^{-\rho a}}{\rho - \mu_i}] \;+\; e^{-\rho a}) f_a(a)\, da \quad (5)$$

Hence the total probability of an individual meeting all criteria at the testing step is:

$$P(\text{positive \& disease endpoint} \mid \text{disease-free}) \;=\; (1 - e^{-3\mu_f})P_2 \;+\; (1 - e^{-3\mu_s})P_2' \;. \quad (6)$$

Note that $P_2'$ is analogous to the expression in equation (5) but with only the numerator $P_1$ replaced by the slow-progressor equivalent (i.e. $p_f \rightarrow p_s$ and $\mu_f \rightarrow \mu_s$) as the denominator remains unchanged.

Finally in the enrollment step, individuals need to become a disease endpoint GIVEN they are disease-free AND were infected in past σ years ('positive'). Again using the definition of conditional probability and the law of total probability, we can express the fast progressor component as:

$$P_3 \;:=\; P(\text{fast} \mid \text{positive \& disease-free}) \;=\; \frac{P(\text{positive \& fast} \mid \text{disease-free})}{P(\text{positive \& fast} \mid \text{disease-free}) + P(\text{positive \& slow} \mid \text{disease-free})} \;. \quad (7)$$

This is simply $\frac{P_2}{P_2 + P_2'}$. Then the total probability of an individual meeting all criteria at the enrollment step is:

$$P(\text{disease endpoint} \mid \text{disease-free \& positive}) \;=\; \frac{(1 - e^{-3\mu_f})P_2 \;+\; (1 - e^{-3\mu_s})P_2'}{P_2 \;+\; P_2'} \;. \quad (8)$$

The expected number needed to contact, test, and enrol (using equations 3, 6, 8) is hence as follows:

$$E[\text{no. contacted to enrol n* disease endpoints}] \;=\; \frac{\text{n*}}{(1 - e^{-3\mu_f})P_1 \;+\; (1 - e^{-3\mu_s})P_1'} \;;$$

$$E[\text{no. tested to enrol n* disease endpoints}] \;=\; \frac{\text{n*}}{(1 - e^{-3\mu_f})P_2 \;+\; (1 - e^{-3\mu_s})P_2'} \;;$$

$$E[\text{no. enrolled to enrol n* disease endpoints}] = \frac{(P_2 + P_2')\text{n}^*}{(1-e^{-3\mu_f})P_2 + (1-e^{-3\mu_s})P_2'} \quad . \text{ (9)}$$

##### Stochastic approach

In the stochastic approach, the time to infection tinf is age-varying and uses a general survival function with empirically-specified hazard rate ρ: $S(a) = \exp(-\int_0^a \rho(u)du)$ for each age a. This expression collapses to the exponential survival function when ρ is constant (as in the analytic approach). For each individual, the time to infection tinf is drawn from this survival distribution with hazard rate ρ equal to the annual force of infection. The time to symptoms tsymp is then calculated as tinf + Exponential($\mu_f$), for rate of progression to disease $\mu_f$ (or $\mu_s$ if a slow progressor).

Once infected, a fraction $p_r$ of individuals are assumed to be 'reverters' who clear infection, with the time at which primary reversion occurs modelled as tinf + Exponential($\mu_r$) for constant annual reversion rate once infected $\mu_r$. There is no reinfection or secondary reversion in the model[2]. We also assume that only one of reversion or disease progression can occur for each individual - the chance of becoming a case vs reverting is hence an exponential race. Individuals are only enrolled if they are still positive (i.e. have not reverted) and are disease-free at the time of recruitment. Once enrolled, individuals can revert or progress to disease *during* the trial, based on their simulated reversion/progression times respectively. The stochastic simulation continues until 50 cases have been enrolled. $p_r$ is taken from recent IGRA/QFT-Plus positivity surveys (Dagnew et al. and Selvaraju et al. [10,11]), while $\mu_r$ is informed by a retrospective study that found the relative increase in IGRA reversion rate to be small after two years of retesting in Germany (Meier & Enders [12]).

#### Model parameters

Parameter values used in our model are shown in Supplementary Table S1. The analytic approach uses a uniform force of infection and uniform probability of being a fast progressor. The stochastic approach additionally explores an age-varying force of infection and age-varying probability of being a fast progressor, and allows primary reversion. Rate of progression to disease as a fast (slow) progressor once infected, $\mu_f$ ($\mu_s$), is assumed to be constant.

| Parameter | Description | Base value (South Africa) | Range explored | Reference |
|---|---|---|---|---|
| minage | Minimum age for trial eligibility | 15 years | - | [3] |
| maxage | Maximum age for trial eligibility | 49 years | - | [3] |
| ρ | Force of infection | 6% for ages 0-4,<br>5% for ages 5-24,<br>7% for ages 25-34,<br>7% for ages 35-49,<br>5% for ages 50+ | 1-10% for ages 0-4, 5-24, 25-34, 35-49, and 50+ | Fitted parameter |
| $p_f$ | Probability of being a fast progressor | 4% for ages 0-10, linearly increasing from ages | 4% for ages 0-10, linearly increasing from ages | [2,13] |

[2]IGRA status is thought to be dynamic in reality and some transient reversion (positive-negative-positive) has been observed in IGRA positivity surveys, but at a low rate over a long (> 6 months) time frame [8,9]. Inclusion of reversion in our stochastic model aims to capture those individuals who clear infection.

| | | | | |
|---|---|---|---|---|
| | | 11-19, 19% for ages 20+† | 11-19, 14% for ages 20+† | |
| $p_s$ | Probability of being a slow progressor | 1 - $p_f$ | 1 - $p_f$ | [2,13] |
| $\mu_f$ | Rate of progression to disease once infected, for fast progressors (annual) | Exponential(1.5) i.e. approx 95% progress within 2 years | - | [1] |
| $\mu_s$ | Rate of progression to disease once infected, for slow progressors (annual) | Exponential(0.003) i.e. approx 4% progress within 15 years | 0.0001-0.00527* | [2,14–20] |
| $p_r$ | Probability of being a reverter once infected | 7.5%‡ | | [10] |
| $\mu_r$ | Rate of primary reversion once infected, for reverters (annual) | Exponential(0.75) i.e. approx 78% revert within 2 years‡ | | [12] |
| σ | Duration of positivity of test | 80 years | 0.5-80 years | [5,6] |
| trial_length | Duration of simulated trial | 3 years | - | [3] |

***Supplementary Table S1*****: Parameters used in our South Africa model.** *The model assumes a high-burden South Africa-like population age distribution. †Probability of being a fast progressor from Vynnycky & Fine is calculated according to age of infection (not current age) [13]. *Range for $\mu_s$ is informed by non-age-varying estimates from the following empirical and modelling studies [2,14–20], excluding those models identified as poorly fitting structures in Menzies et al. [21]. ‡Probability of primary reversion once infected is informed by Dagnew et al. Table S6 but rescaled by number of Day 1 positives and the percentage of individuals who completed the study (i.e. reported a Month 12 IGRA status) [10]. This yields an approximate probability of primary reversion for those already positive, rather than probability of primary reversion among all participants as reported. For those individuals who are reverters (7.5%), we assume a constant reversion rate of 0.75 to give approximately 78% reverting within two years of infection, motivated by Meier & Enders [12].*

As with all modelling studies, our analysis is subject to various limitations. We assume perfect test sensitivity and specificity, there is no loss-to-followup, and we do not explicitly model co-infection with HIV. Force of infection is also highly uncertain given the available data.

#### Cost estimates

Costings are difficult to estimate but for the purposes of this simple analysis, we assume a single long-term positivity test such as IGRA costing on the order of $150 in 2026 US dollars, and trial participation for a single enrolled individual costing on the order of $23,000. A recent systematic review of infection test costs estimated IGRA tests to cost approximately $108.22 ($27.32-$304.56) when scaled up to 2026 US dollars [22]. The review included two studies from South Africa both on the higher end of this range but note that all reported economic costs included both the costs of the tests (disposables, administration, reading, lab technicians) and additional screening costs (two clinic visits, one chest radiograph). We hence believe $150 to be a reasonable estimate for unit IGRA-style test costs. Trial participation costs were informed by the M72/AS01E phase III trial's 2023 announcement of $550 million total cost for 26,000 planned enrolled participants, equivalent to approximately $603 million in 2026 US dollars [23]. This suggests a crude unit cost of approximately $23,200 per enrolled participant in 2026. Hence for this simple analysis, we assume trial participation for a single enrolled individual to be at least 150 times more expensive than a single long-term positivity infection test. Costings described here are highly uncertain and vary by location; as such, our results present only the *relative* cost of test vs trial enrolment, not absolute values.

### Model fitting and validation procedures

#### Analytic and stochastic methods

As a first sanity check, we confirmed that the analytic and stochastic approaches aligned when run with uniform age structure and consistent model parameters (Supplementary Figure S2).

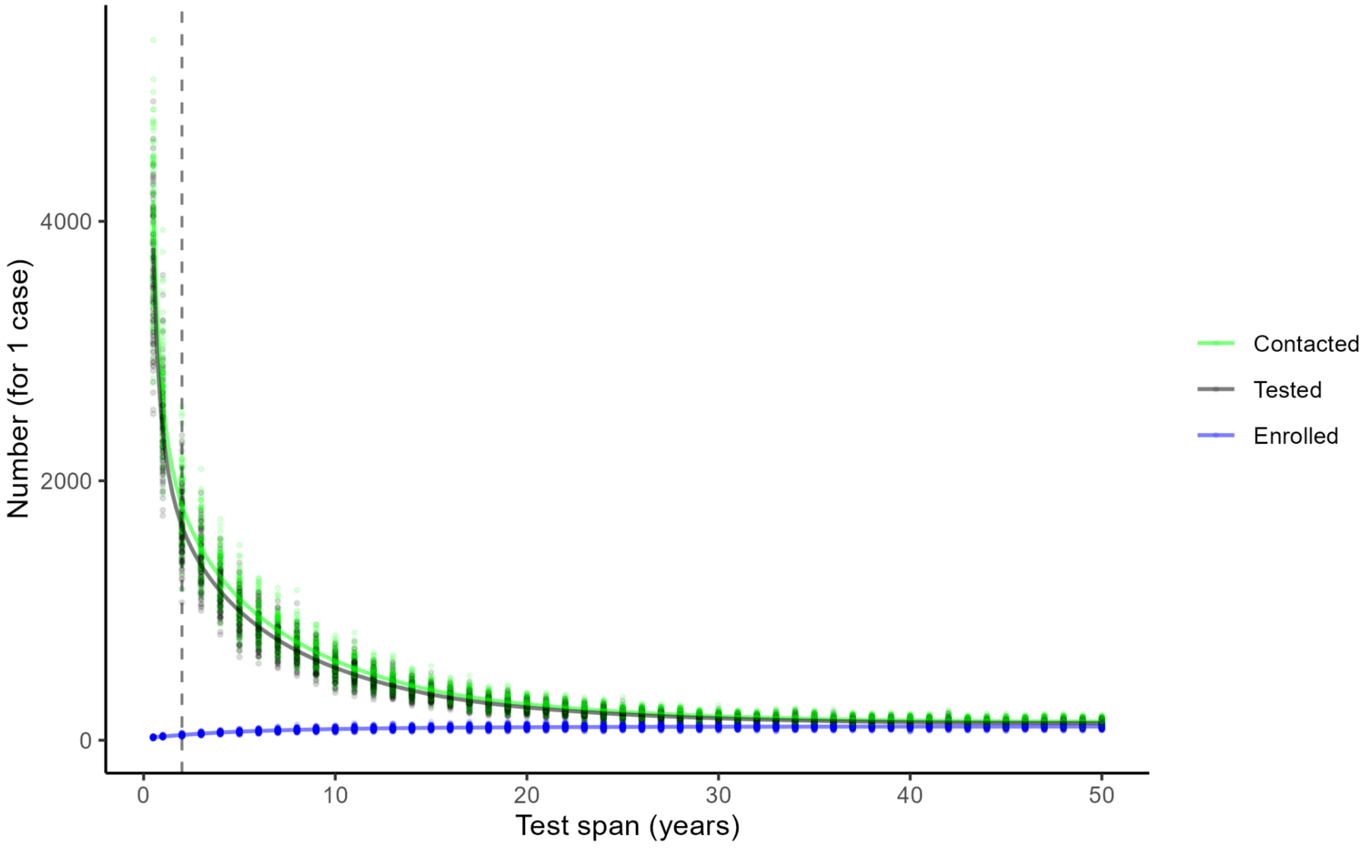


***Supplementary Figure S2: Plot comparing the stochastic (points) and analytic (curve) approaches, for a South Africa-like population with uniform force of infection and probability of being a fast progressor (6% and 5% respectively) and no reversion.***

#### Force of infection

Age-varying force of infection, ρ, was chosen to emulate literature estimates of IGRA positivity in a high burden setting in South Africa (Table S6 in Dagnew et al. [10]). For a simple catalytic model with hazard rate ρ, we would expect ρ around 5% to emulate the IGRA positivity observed in Dagnew et al. [24]. A 2010 study on Tuberculin Skin Test positivity by age further informed using a piecewise-constant force of infection, with differing values for ages 0-4y, 5-24y, 25-34y, 35-49y, and 50y+ [25]. We ran the model with the force of infection varying from 1-10%, with 25,040 parameter combinations each run for 5 stochastic repetitions. Parameter combinations were chosen to best match the Dagnew et al. estimates of IGRA positivity by age in South Africa, as well as wider literature estimates of annual risk of Tuberculosis infection (ARTI) and annual case data [10,25,26]. 523 parameter combinations captured the range observed in the literature after 5 stochastic repetitions. The most plausible of these combinations and those with near-misses of the threshold bounds were rerun for 25 and then 100 stochastic repetitions to confirm performance. From these candidates, we finally selected a force of infection of 6% for ages 0-4, 5% for ages 5-24, 7% for ages 25-34, and 7% for ages 35-49, and 5% for ages 50+ as it gave estimates for IGRA positivity closest to the literature with non-maximal $\mu_s$ and mirrored detailed TST positivity by age trends from Wood et al.. Full results of the model fitting are shown in Supplementary Figure S3.

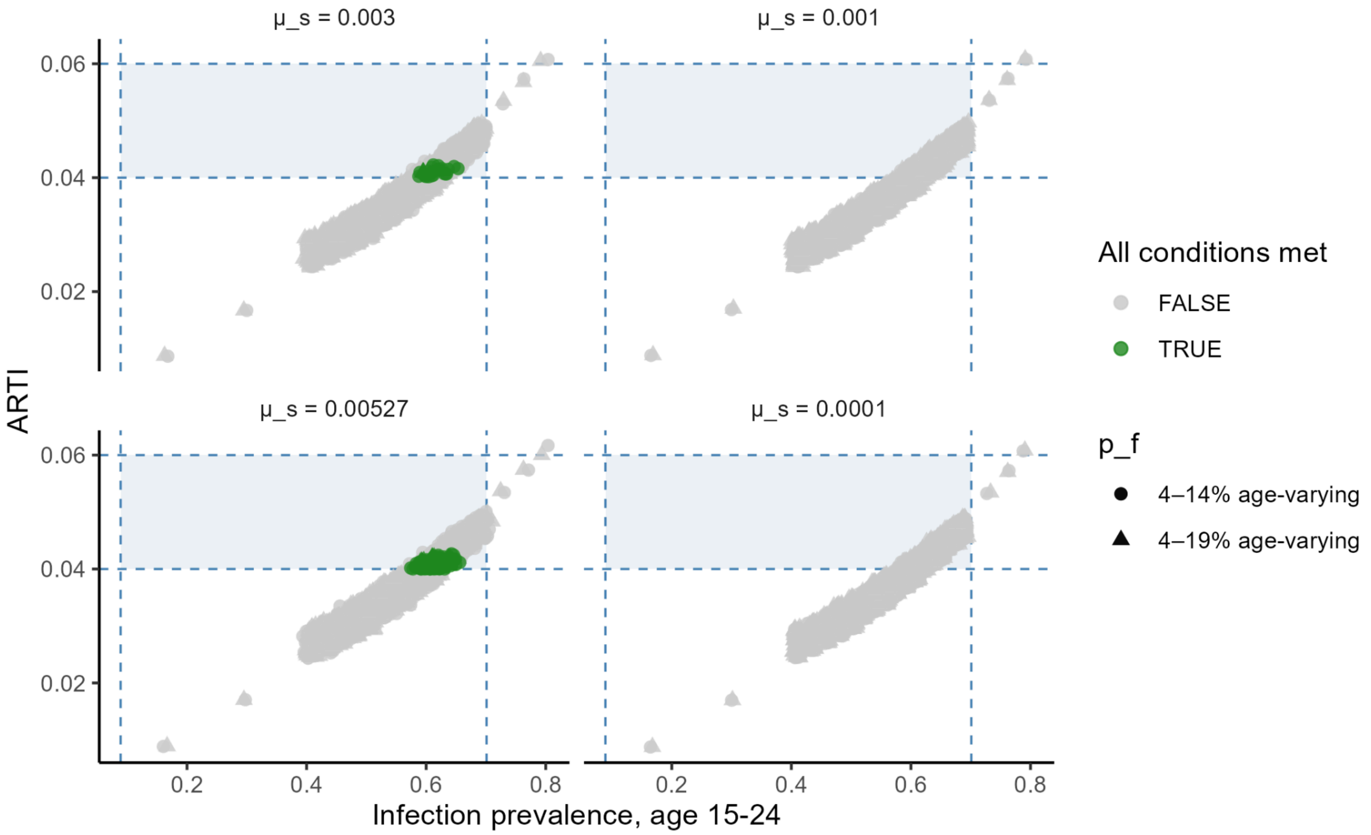


***Supplementary Figure S3:* Results from fitting the age-varying force of infection, ρ, to data on IGRA positivity by age, annual risk of Tuberculosis infection (ARTI), and case incidence.** *A range of values for ρ, $p_f$, $\mu_s$ were explored. Points represent different values of ρ, with $p_f$ depicted by point shape (circle/triangle) and $\mu_s$ depicted by facet (0.0001, 0.001, 0.003, 0.00527). All other model parameters used the base values from Supplementary Table S1. The target ranges for infection prevalence at age 15-24 and ARTI are shown (blue shaded regions). ARTI is calculated as 1- (1 - P(a))$^{1/a}$ for infection prevalence P and age a. The 'all conditions met' marker (green) identifies those parameter combinations that met all of the following criteria: median infection prevalence of 8.9-70.1% at ages 15-24 and 19.6-75.1% at ages 25-34; median ARTI of 4-6%; and median cases per 100k per year of 242-571. 523 of the 25,040 parameter combinations considered met all conditions after 5 stochastic repetitions - the most plausible of these candidates and other near-misses in the search parameter neighbourhood were rerun for 25 and then 100 repetitions to confirm performance. Note that no scenarios with $\mu_s$ = 0.0001 or 0.001 met all conditions.*

#### Trial emulation

We additionally used our model to emulate results of the recently-completed M72/AS01E phase IIb trial by Tait et al. [3]. This trial enrolled 3289 participants in the according-to-protocol efficacy cohort and reported 26 cases in the placebo arm and 13 cases in the vaccine arm over the three-year duration of the trial. In the absence of vaccination, we would therefore expect approximately 52 (26+26) potential cases among the 3289 participants during the trial. Using the fitted force of infection ρ, our model recorded a median case count within 20% of the 52 target from 3289 enrolled participants if $\mu_s$ was large (i.e. $\mu_s$=0.00527), OR if $\mu_s$ = 0.003 and $p_f \geq$ 30%. These results are from 100 stochastic repetitions per parameter set. When $\mu_s$=0.00527 was used, both of the $p_f$ variations from Supplementary Table S1 (age-varying 4-14%, age-varying 4-19%) achieved the target case count, as did uniform $p_f$ in 5% increments from 5-25%.

#### Model scenarios

The following baseline and sensitivity scenarios were explored (Supplementary Table S2):

| Scenario | | Details |
|---|---|---|
| 1 | Test-based enrolment in a high-TB country | This is our main scenario which uses a high burden South Africa-like population. Parameter values are as defined in Supplementary Table S1. |
| 2 | Test-based enrolment with a varied vaccine trial site | Same as 1) but using an India-like population (low ARTI) and Kenya-like population (medium ARTI) as the age distribution. The force of infection for each is fitted to data on IGRA positivity by age, ARTI, and cases following the same approach as was done for South Africa [10,11,29–31]. $p_r$ is set to 16.1% for Kenya and 20.6% for India* [10]. A range of $p_f$ values are considered. All other model parameters take the values in Supplementary Table S1. |
| 3 | Non-test based enrolment with no age restrictions | A high burden South Africa-like population and individuals are enrolled immediately provided they are disease-free (regardless of whether or not they are infected). minage=15; maxage=49. |
| 4 | Non-test based enrolment - young people | As 3) but a lower age range is considered: minage=12; maxage=34. A trial of this type would aim to capture peak TB incidence in adolescents in high burden countries [27]. |
| 5 | Non-test based enrolment - household contacts of recent cases | As 3) but individuals experience a large instantaneous force of infection at the time of recruitment if not previously infected, modelled as a 26% chance of successful household infection at the current age following a recent household contacts trial in India [28]. If previously infected, there is no change to an individual's probability or rate of progression. Here minage=6 and maxage=99. |

***Supplementary Table S2: A description of model scenarios.*** **The probability of primary reversion once infected, $p_r$, for Kenya is informed by Dagnew et al. [10]. For India, Dagnew et al. reported an unusually low number of positives in its single site so $p_r$ is instead informed by the equivalent data for Bangladesh - a country with similar TB incidence and risk factors - and IGRA positivity bounds are taken from Selvaraju et al. [11,32].*

### Supplementary results

#### Test-based enrolment (varied trial sites)

Results for a test-based recruitment with varying duration of positivity in a high-burden South Africa-like population are shown in Supplementary Figures S4 and S5. We see that the number needed to enrol for a sufficiently powered trial could be significantly reduced if a test for recent infection ($x < 5$ years) is used. We additionally explored varied vaccine trial sites, using a Kenya-like population and an India-like population (Supplementary Table S3) [4]. For these analyses, force of infection $\rho$ was refitted following the same procedure as South Africa, again chosen to best match data on IGRA (or QFT) positivity, ARTI, and case incidence [10,11,29–31]. $p_f$ was varied to capture the differences in relevant risk factors between countries (differing prevalence of HIV, malnutrition, diabetes).

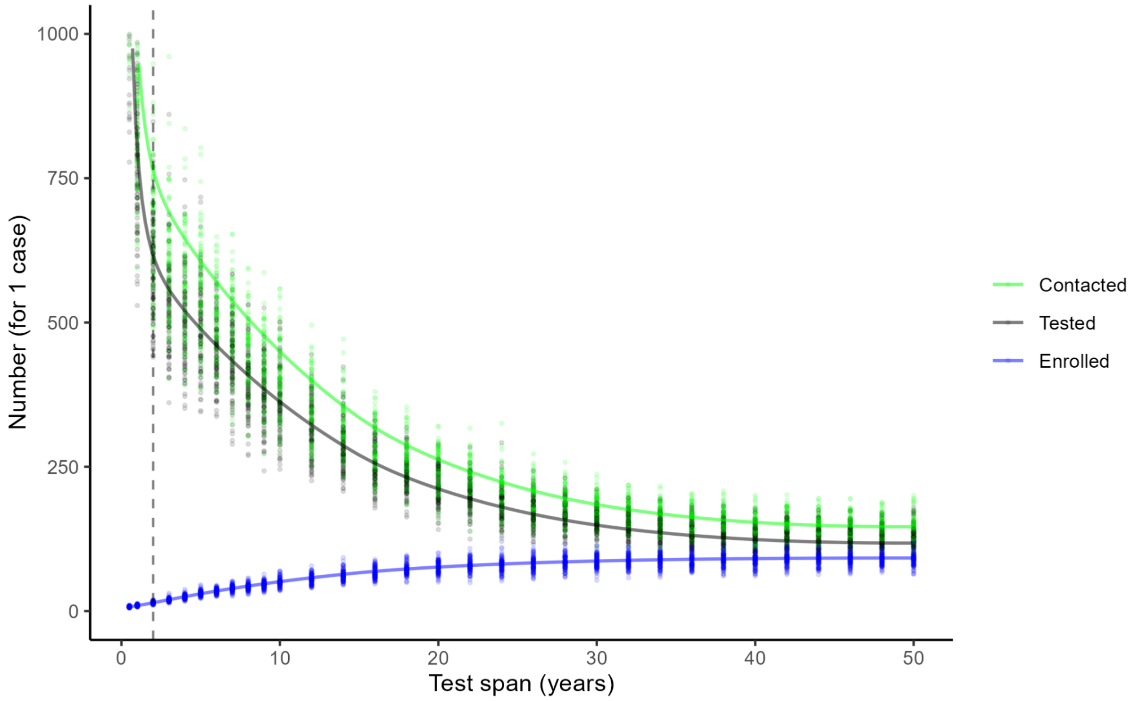


***Supplementary Figure S4*: Modelling estimates of the number of individuals that need to be contacted, tested, and enrolled in TB vaccine trials to observe one disease endpoint in a high burden South Africa-like population.** *The x-axis depicts duration of positivity of the test used for recruitment, ranging from very short (0-2 years) akin to a test of recent infection to extended positivity akin to an IGRA. Both the stochastic (points, 100 repetitions) and analytic (curve) approaches are shown.*

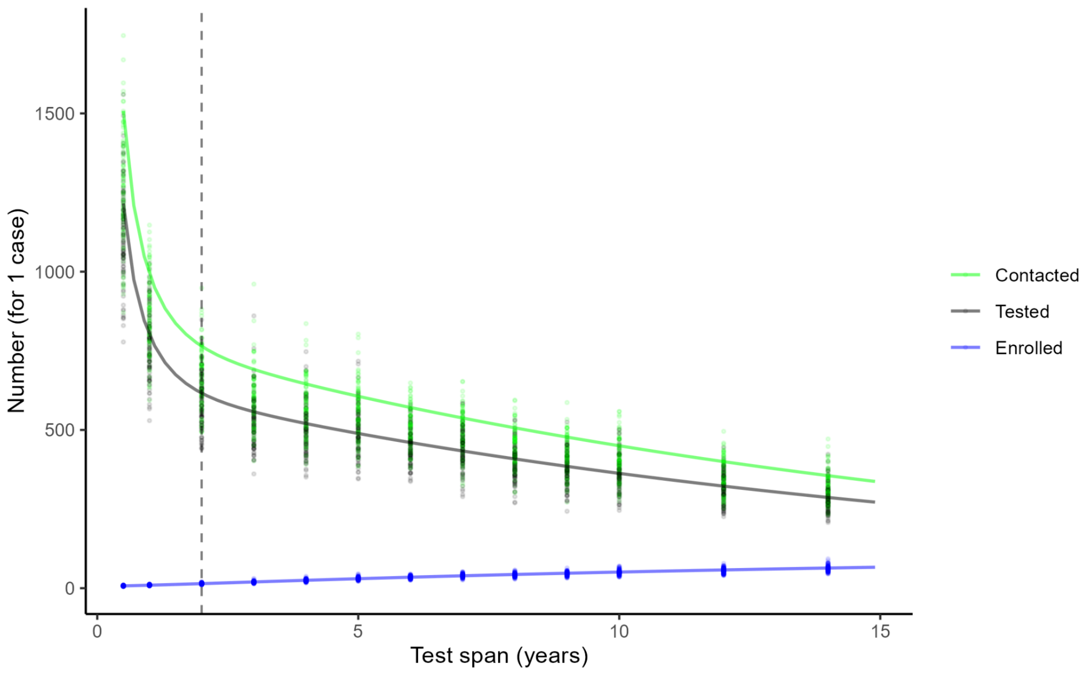


***Supplementary Figure S5*: As *Supplementary Figure S4* but focusing in on the 0-15 year test span results.** *See Supplementary Figure S4 legend.*

| Country | Literature estimates | | | | Model parameters | | Model estimates from trial population aged 15-49y | | | | All conditions met? |
|---|---|---|---|---|---|---|---|---|---|---|---|
| | IGRA/QFT positivity | | ARTI | Case incidence per 100k per year | ρ ages 0-4, 5-24, 25-34, 35-49, 50+ | $p_f$ | Infection prevalence | | ARTI | Case incidence per 100k per year | |
| | ages 15-24 | ages 25-34 | | | | | ages 15-24 | ages 25-34 | | | |
| India | 0.098-0.158 | 0.171-0.252 | 0.00-0.02 | 160-218 | 2,1,3,1,1% | 4-14% | 0.176 | 0.286 | 0.011 | 101 | FALSE |
| | | | | | | 5% | 0.181 | 0.288 | 0.011 | 85 | FALSE |
| Kenya | 0.182-0.500 | 0.256-0.659 | 0.016-0.036 | 109-339 | 5,2,5,4,2% | 4-14% | 0.349 | 0.490 | 0.020 | 157 | TRUE |
| | | | | | | 5% | 0.351 | 0.487 | 0.020 | 161 | TRUE |
| South Africa | 0.089-0.701 | 0.196-0.751 | 0.04-0.06 | 242-571 | 6,5,7,7,5% | 4-19% | 0.595 | 0.742 | 0.041 | 243 | TRUE |

***Supplementary Table S3*: Median infection prevalence, ARTI, and case incidence estimates from model scenarios of varied trial sites, compared to best estimates from the literature.** *Results are from 100 stochastic repetitions, using the best-choice age-varying force of infection from model fitting. Where model estimates met the literature ranges, the cell is coloured green. The 'conditions met' column displays those parameter sets where the model gave plausible outputs for all of infection prevalence by age, ARTI, and case incidence [10,11,29–31]. The most plausible force of infection ρ and probability of being a fast progressor $p_f$ values are shown (range explored: 1-10% for ρ; 5%, 4-14% age-varying, and 4-19% age-varying for $p_f$). Other parameter values are as described in Supplementary Table S2.*

From Supplementary Table S3, we saw that using an age-varying force of infection of 2-5% for Kenya and 1-3% for India could be plausible, although noting that case incidence in India was underestimated and infection prevalence overestimated in our model. Some ARTI estimates were slightly low but this is to be expected due to our method of calculation [9].

Results of the number needed to test and enrol for each country are shown in Supplementary Figure S6 and Supplementary Figure S7. We observed a significant increase in the number of individuals that would need to be enrolled per case as duration of positivity of test increased, from approximately 15-20 at σ=2 to 80-90 at σ=50+ in these model scenarios. In other words, model estimates suggest a test of recent infection with say σ=1-2 years could significantly reduce the required enrolment size of TB vaccine trials by up to six-fold. The number of tests needed, however, would increase significantly.

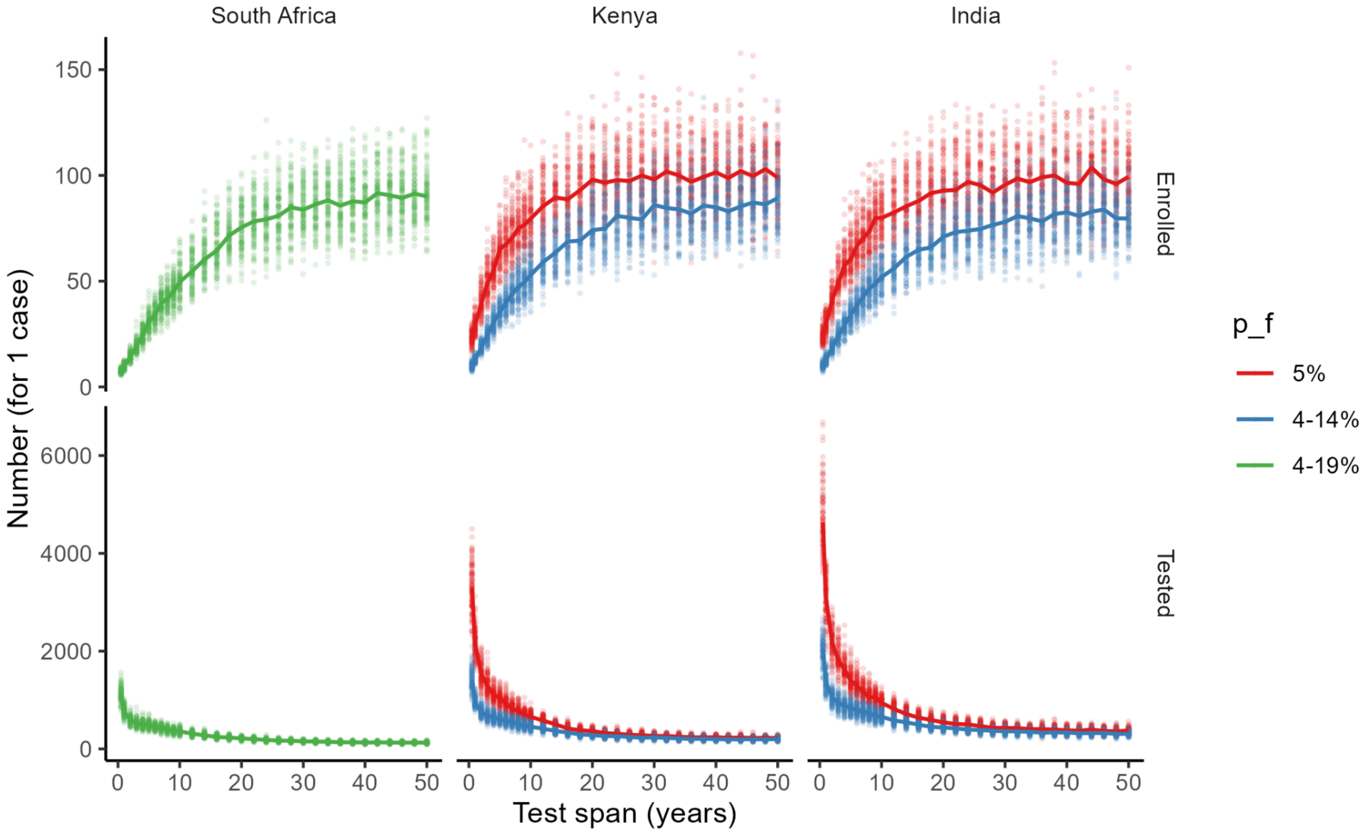


***Supplementary Figure S6*: Model results for test-based scenarios in varied trial sites (South Africa, India, Kenya).** *The expected number of people needed to test and enrol are shown, for a range of test duration of positivity/ test span σ. Probability of being a fast progressor* $p_f$ *is shown as linetype, using the most plausible parameter combinations reported in Supplementary Table S3. Plots show results from 100 repetitions of the stochastic model approach (points). Here lines represent the median of all 100 repetitions at each test span. Analytic approach is not shown.*

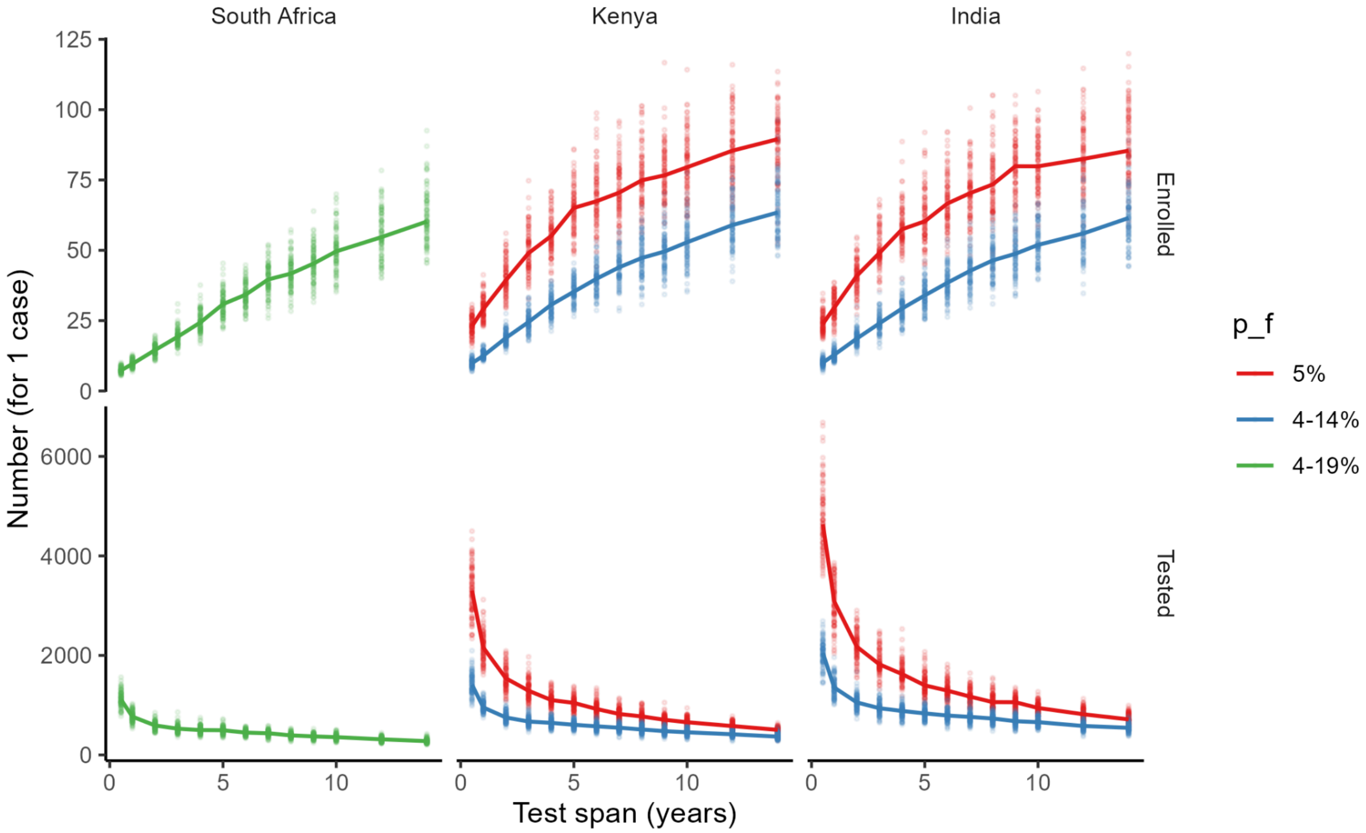

***Supplementary Figure S7*: As Supplementary Figure S6 but focusing in on the 0-15 year test span results.** *See Supplementary Figure S6 legend for details.*

##### Economic impact

We additionally estimated the potential costs saved when recruiting with a test of recent infection (akin to TB-TASA) compared to the current IGRA. Model estimates for number needed to test and enrol were taken from the simulations depicted in Supplementary Figure S6 and Supplementary Figure S8. Although a test for recent infection could enable significantly smaller trials, more individuals would need to be tested. In Supplementary Table S4, we see that costs have the potential to be cheaper provided new recency tests are not more than 5-10 times more costly than current longer-term positivity infection tests.

| Country | Model parameters | | Model estimates | | | | Estimated costs for σ=2 | | |
|---|---|---|---|---|---|---|---|---|---|
| | ρ ages 0-4, 5-24, 25-34, 35-49, 50+ | $p_f$ | No. needed to test median [IQR] | | No. needed to enrol median [IQR] | | Expected cost of tests (relative to σ=50) | Expected cost of trial participation (relative to σ=50) | **Recency test cheaper overall if:** (zero cost difference ratio) |
| | | | σ=2 years | σ=50 years | σ=2 years | σ=50 years | | | |
| India | 2,1,3,1,1% | 4-14% | 1055 [958-1153] | 309 [282-337] | 19 [17-20] | 80 [74-87] | 3.4X | 0.24Y | 0.24*150/3.4 = 10.6 |
| | | 5% | 2174 [2001-2381] | 375 [341-407] | 41 [37-44] | 99 [91-109] | 5.8X | 0.41Y | 0.41*150/5.8 = 10.6 |
| Kenya | 5,2,5,4,2% | 4-14% | 752 [692-834] | 202 [180-216] | 19 [17-21] | 89 [79-96] | 3.7X | 0.21Y | 0.21*150/3.7 = 8.5 |
| | | 5% | 1539 [1388-1717] | 221 [206-247] | 39 [36-44] | 99 [92-110] | 7.0X | 0.40Y | 0.40*150/7.0 = 8.6 |
| South Africa | 6,5,7,7,5% | 4-19% | 592 [530-649] | 128 [118-137] | 15 [13-16] | 90 [83-96] | 4.6X | 0.17Y | 0.17*150/4.6 = 5.5 |

***Supplementary Table S4*: Potential costs saved if TB vaccine trial recruitment could utilise a test for recent infection compared to current IGRA recruitment procedures.** *Model estimates are the median number needed to test/enrol for 1 disease endpoint, across 100 stochastic repetitions, with the interquartile range shown in square brackets. Cost calculations assume a relative unit cost (combining equipment, procurement, and delivery) of X per TASA-style test (σ=2) compared to an IGRA-style test (σ=50+), and a relative unit cost of Y=150 per individual enrolled in a three-year trial compared to one IGRA-style test.*

#### Non-test based enrolment

Lastly, we used our model to explore non-test-based enrolment. Scenarios are as described in Supplementary Table S2 and results are shown in Supplementary Figure S8 and Supplementary Table S5. Overall, non-test-based scenarios are expected to require a larger trial size, which is more variable by setting, unless recruitment is focused on household contacts of recent cases. In the household contacts scenario, under the assumption that contacts have a 26% chance of being infected at the time of enrollment [28], we estimate that approximately 2,750-3,350 individuals would need to be enrolled for a powered trial (50 disease endpoints) in a South Africa-like population, compared to approximately 4,150-4,800 individuals in the extended IGRA-test based scenarios above.

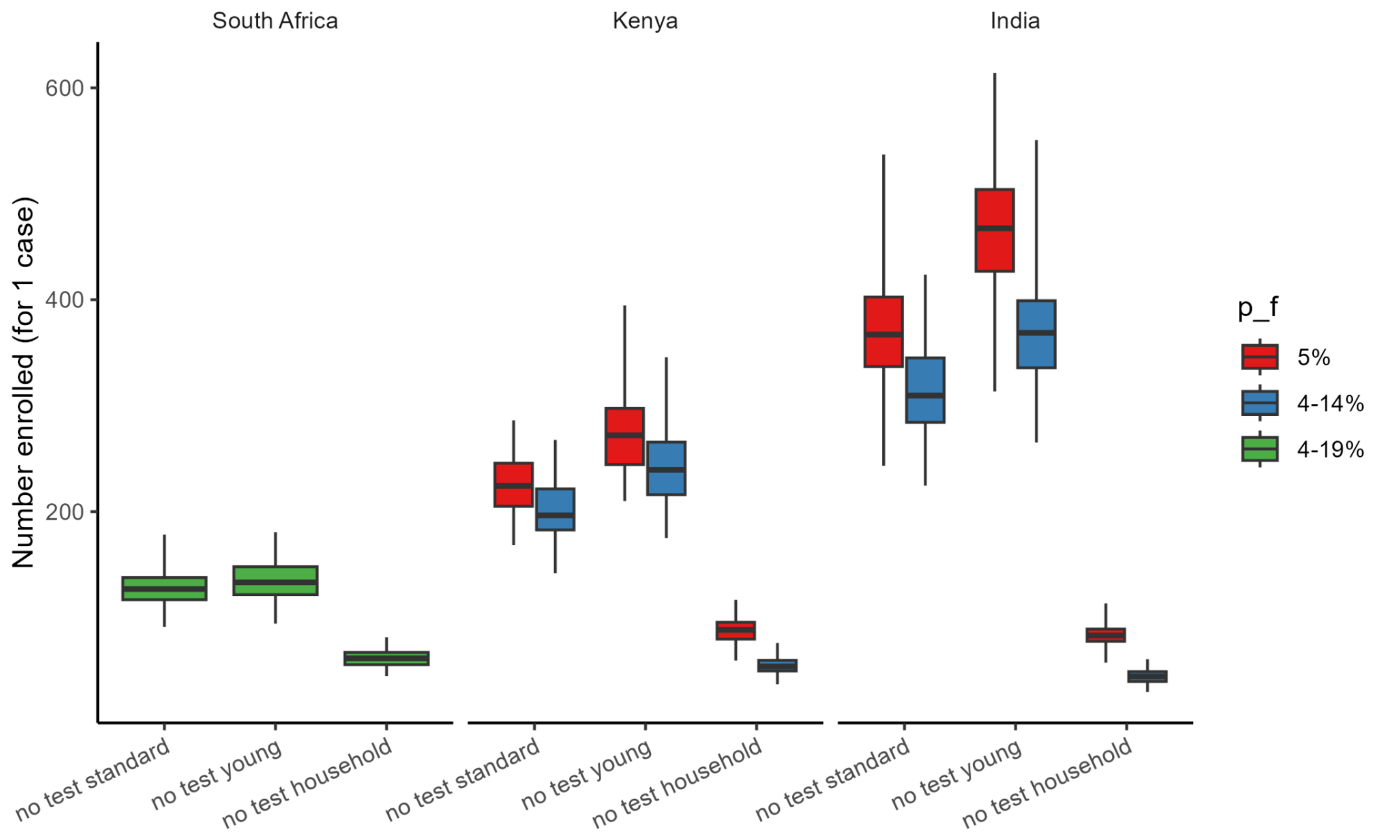


***Supplementary Figure S8: Model results for non-test based scenarios (standard ages 15-49y; younger ages 12-34y; household contacts of recent cases).*** *South Africa, India, and Kenya are shown as facets, with probability of being a fast progressor, $p_f$, values shown as colours. See Supplementary Table S2 for a description of each scenario and Supplementary Table S3 for parameter values used. Plots show results from 100 repetitions of the stochastic model approach.*

| Country | Model parameters | | Model estimates | | | | | |
|---|---|---|---|---|---|---|---|---|
| | ρ ages 0-4, 5-24, 25-34, 35-49, 50+ | $p_f$ | No. needed to contact median [IQR] | | | No. needed to enrol median [IQR] | | |
| | | | No test - standard | No test - young | No test - household | No test - standard | No test - young | No test - household |
| India | 2,1,3,1,1% | 4-14% | 322 [297-360] | 378 [344-409] | 47 [42-51] | 310 [284-345] | 369 [336-399] | 44 [40-49] |
| | | 5% | 378 [347-415] | 477 [435-514] | 86 [80-92] | 367 [337-403] | 467 [427-504] | 83 [78-89] |
| Kenya | 5,2,5,4,2% | 4-14% | 210 [194-236] | 249 [225-277] | 57 [53-63] | 196 [183-221] | 239 [216-266] | 54 [50-60] |
| | | 5% | 236 [216-259] | 283 [254-309] | 93 [84-100] | 224 [205-246] | 272 [244-298] | 88 [80-96] |
| South Africa | 6,5,7,7,5% | 4-19% | 141 [131-154] | 144 [131-160] | 69 [63-75] | 127 [117-138] | 133 [122-148] | 62 [55-67] |

***Supplementary Table S5: Model estimates corresponding to Supplementary Figure S8.*** *Table shows the results for non-test based scenarios (standard ages 15-49y; younger ages 12-34y; household contacts of recent cases). Values represent the median number needed to contact/enrol for 1 disease endpoint, across 100 stochastic repetitions, with the interquartile range shown in square brackets.*

We can also compare promising non-test-based scenarios such as household recruitment to the test-based scenarios, as in Supplementary Figure S9. Here the equivalence point for number needed to enrol for household vs test-based scenarios is shown as dashed lines. Model findings suggest that household recruitment would be approximately equivalent (in terms of required trial size) to a test with 8-15 years DOP.

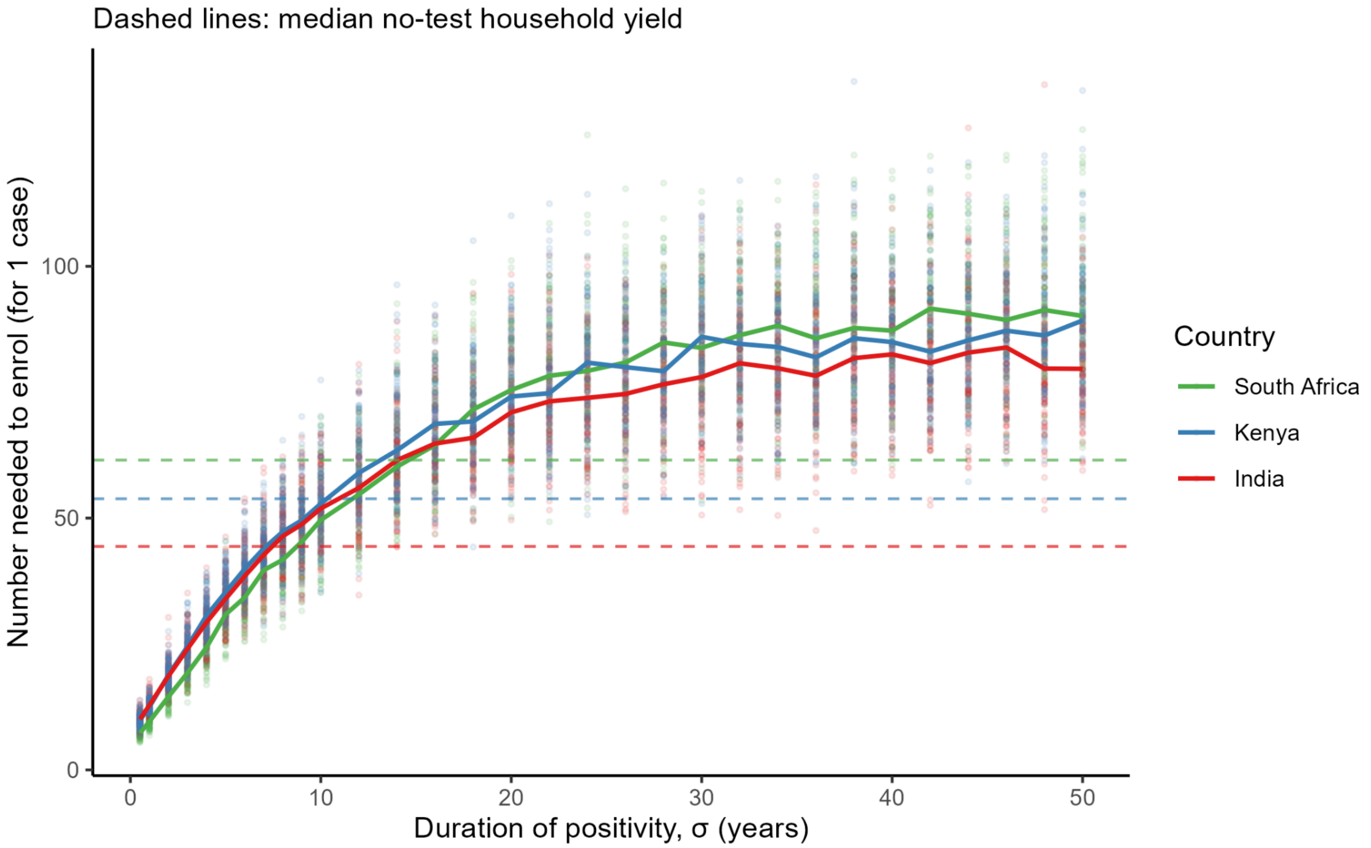


***Supplementary Figure S9:* Comparison of test-based scenarios (x-axis) against household recruitment (dashed lines).** *The intersection point represents at what duration of positivity test-based recruitment would need similar trial sizes to household recruitment.*

#### Sensitivity analysis

We additionally explored the effect of trial size on observed stochasticity. Supplementary Figure S10 considers trial simulations continually recruiting until 5, 50, and 500 cases have been enrolled. We see that observed stochasticity is related to the size of the trial itself, as expected.

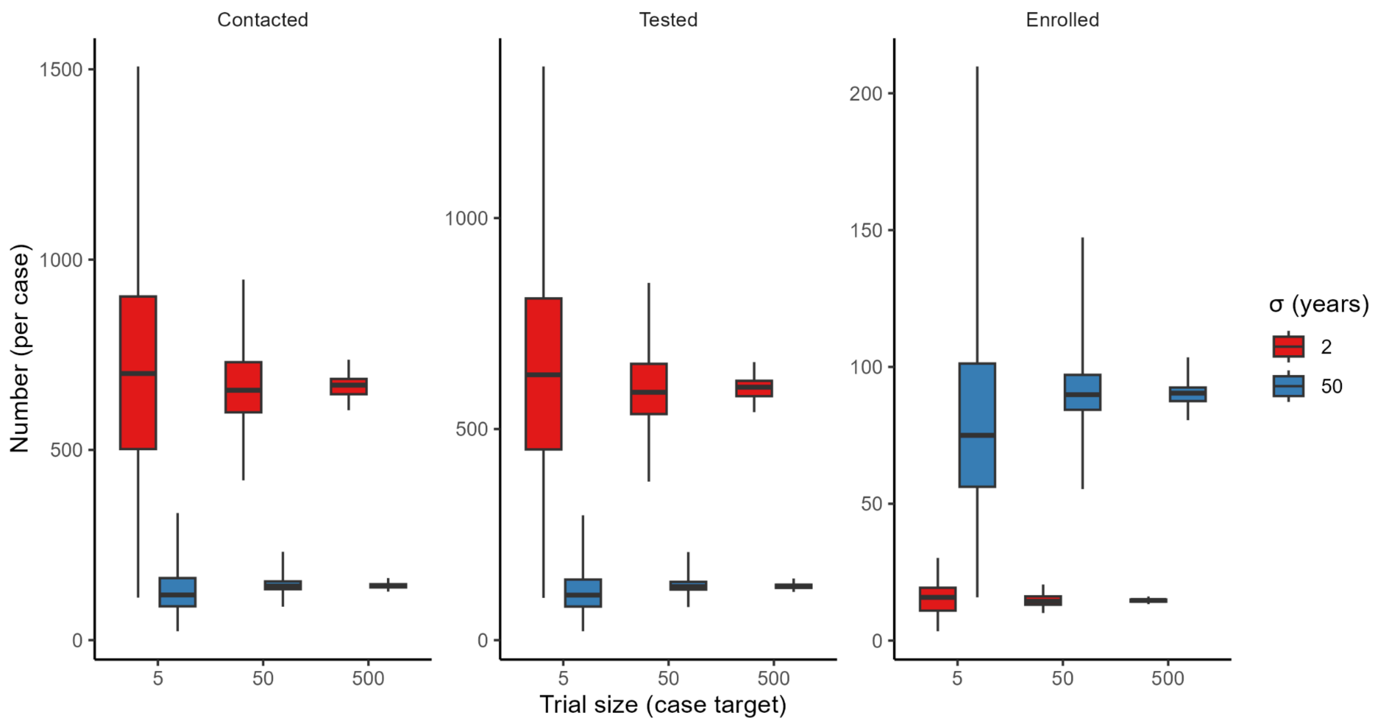


***Supplementary Figure S10*: Exploring the effect of trial size on observed stochasticity, for the South Africa model.** *Simulations are run for 100 stochastic repetitions and until either 5, 50, or 500 cases are enrolled respectively.*

Lastly we examined sensitivity of parameter choices on model outputs (number needed to contact, test, and enrol). Five parameter values were varied by ± 20% from their base value in Supplementary Table S1 for the high burden South Africa-like population. Results are shown in Supplementary Figures S11, S12, and S13. From Supplementary Figure S11, we see the effect of perturbing parameter values on the number needed to contact, test, and enrol. For tests of recent infection, parameters $p_f$ and $\mu_f$ - the probability of being a fast progressor and the rate of progression to disease for fast progressors - are expected to have the largest effect on model outcomes with a 20-30% change from baseline. For extended-posivity (IGRA) tests on the other hand, misspecifying parameter $\mu_s$ is likely to affect model outcomes most. Supplementary Figure S12 shows if parameters affect the *ratio* of contacted, tested, or enrolled (minimal), and finally Supplementary Figure S13 examines if the relative performance of IGRA vs recency tests holds up under parameter perturbation. For example, on the left, each bar aims to show how much perturbing each parameter shifts the σ=0.5-vs-σ=50 enrolment (or test) ratio. Depending on the choice of parameter values, there may be an up to 30% change in the relative number needed to test and enrol when using a test for recent infection versus an IGRA. Overall, our results are most sensitive to parameter $p_f$.

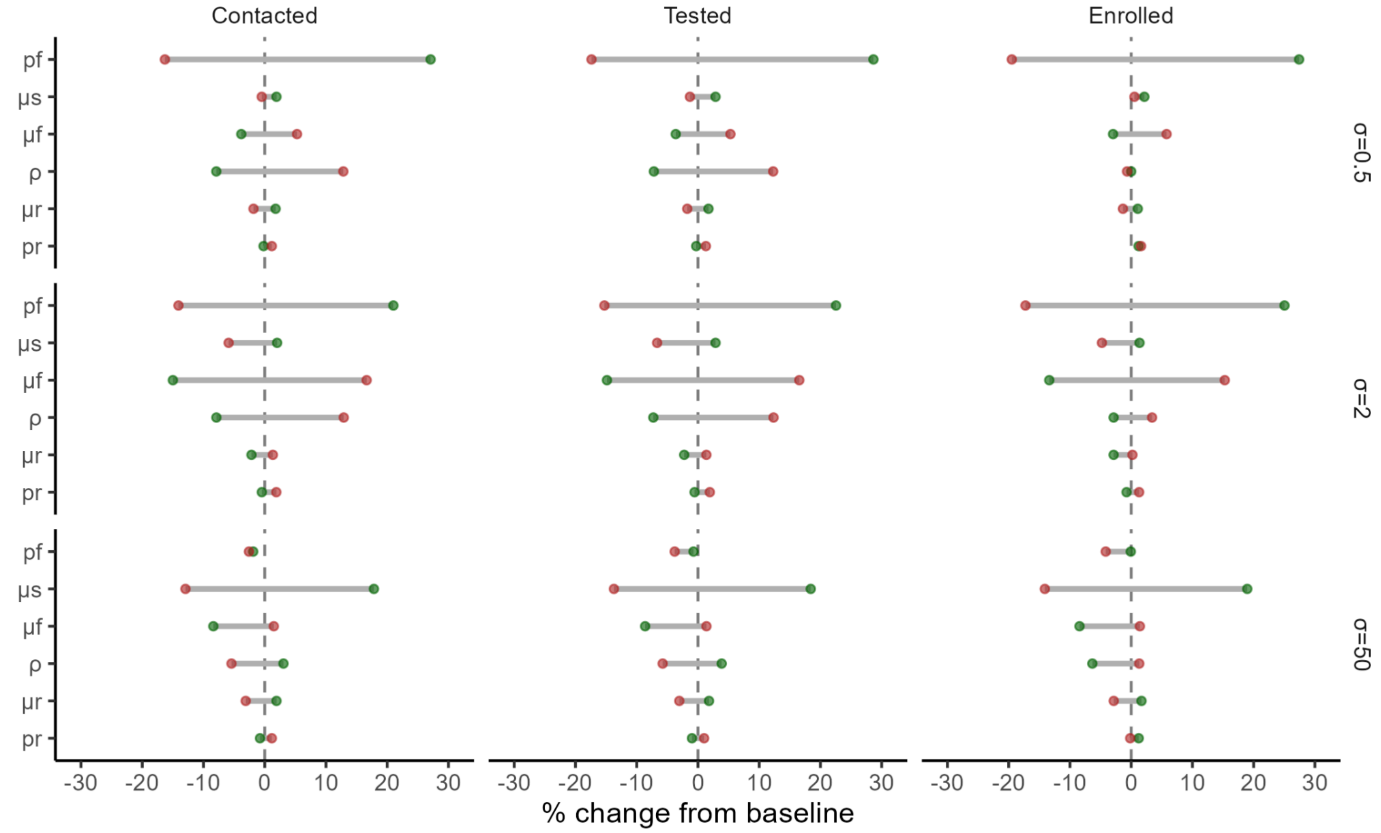


***Supplementary Figure S11*: Exploring the sensitivity of parameter values to model outputs, for the South Africa model.** *Key parameter values ($p_f$, $\mu_s$, $\mu_f$, ρ, $\mu_r$, $p_r$) were varied by ± 20% (green = positive; red = negative). The effect on the number needed to contact, test, and enrol is shown, for different test spans σ. Results are from 100 stochastic repetitions.*

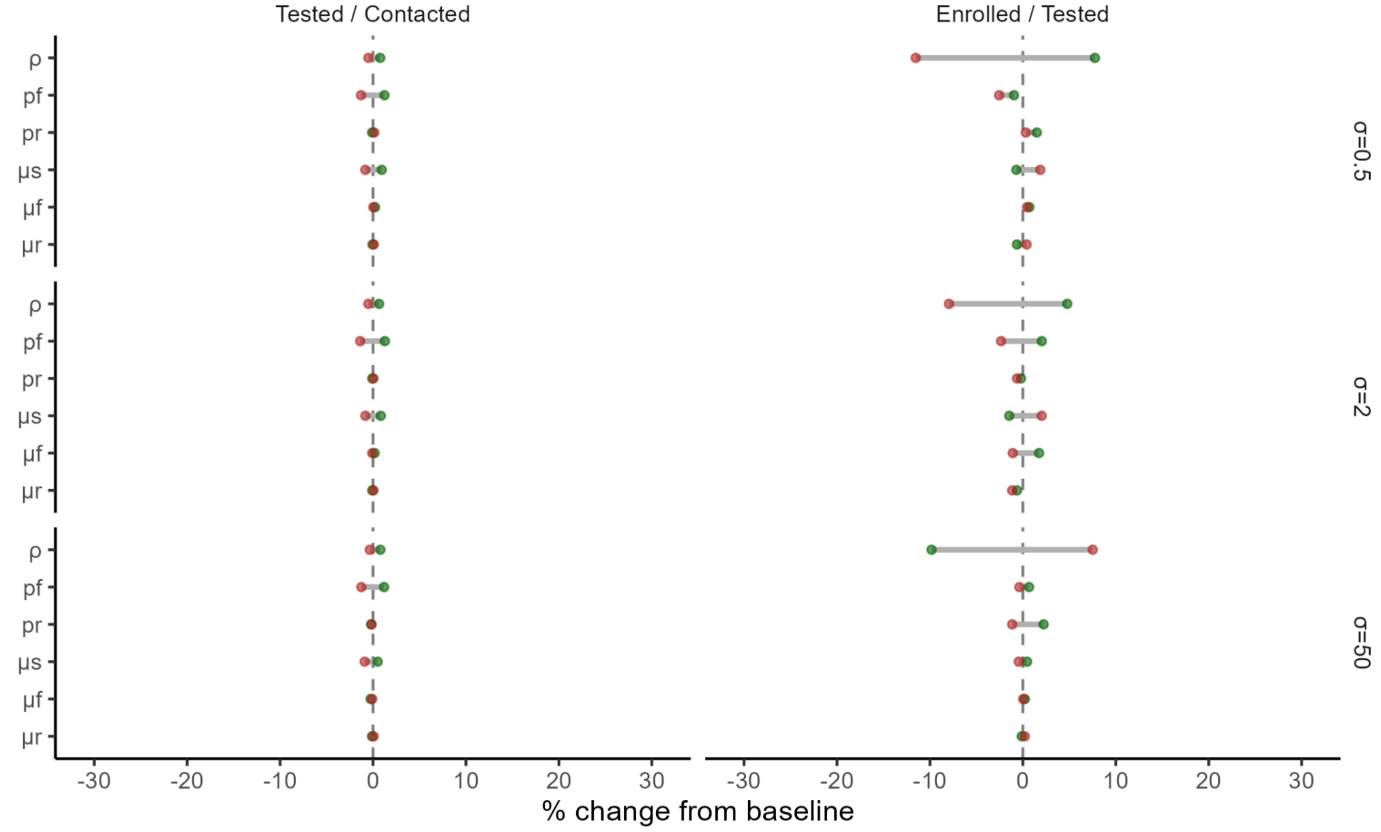


***Supplementary Figure S12*: Exploring the sensitivity of parameter values to relative model outputs, for the South Africa model.** *Key parameter values ($p_f$, $\mu_s$, $\mu_f$, ρ, $\mu_r$, $p_r$) were varied by ± 20% (green = positive; red = negative). The relative effect on the number needed to test versus contact, and enrol versus contact is shown. Results are from 100 stochastic repetitions.*

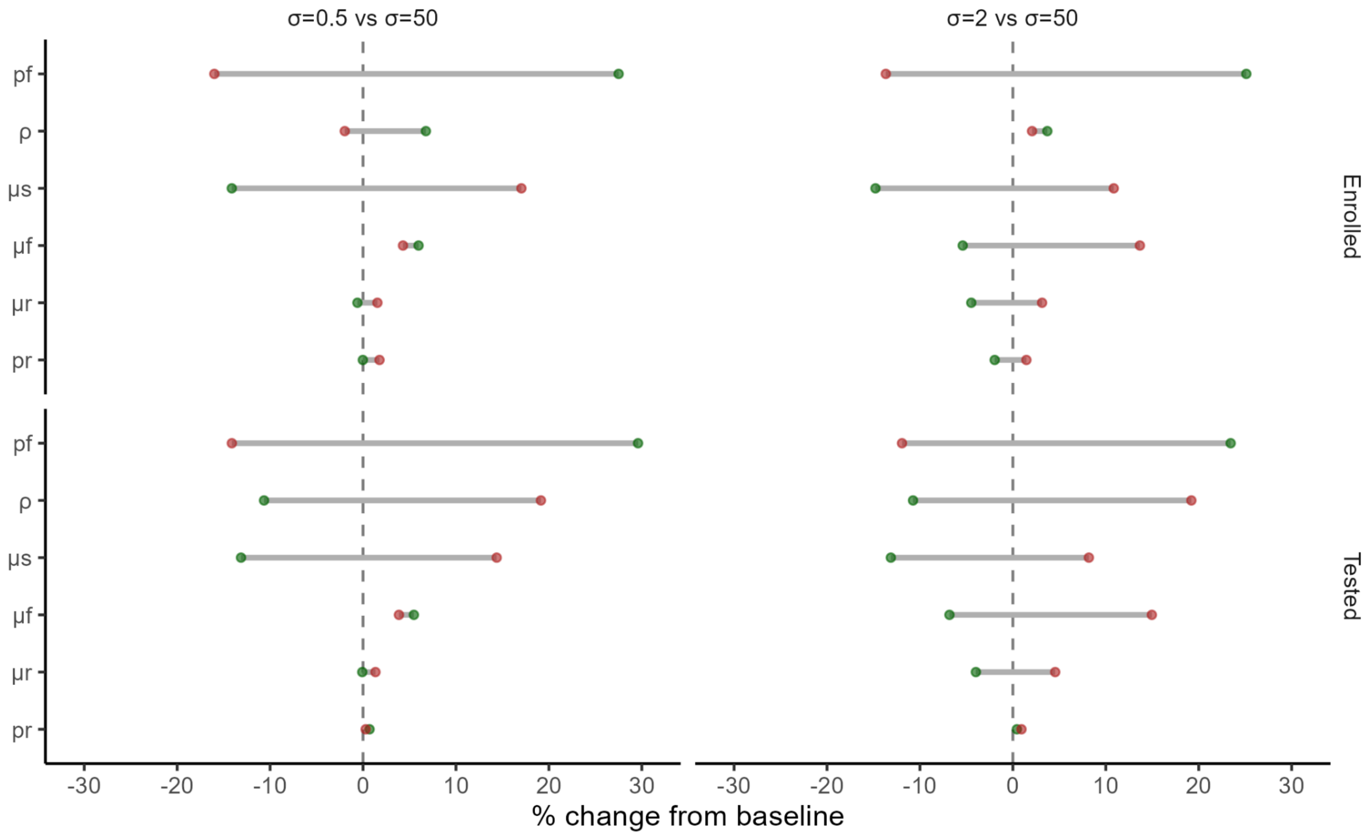


***Supplementary Figure S13: Exploring the sensitivity of parameter values to the relative efficiency of different recruitment strategies.*** *Key parameter values ($p_f$, $\mu_s$, $\mu_f$, $\rho$, $\mu_r$, $p_r$) were varied by ± 20% (green = positive; red = negative), for the South Africa model. The effect on the number needed to enrol and test for a 6-months versus extended positivity test, and a 2 years versus extended positivity test is shown. Results are from 100 stochastic repetitions.*
